# Vaccination recommendations to others among physicians and the general public: effects of birth-year-based vaccination policy changes assessed by regression discontinuity analysis

**DOI:** 10.64898/2026.07.15.26358203

**Authors:** Michio Murakami, Hiroki Kato, Fumio Ohtake

## Abstract

**Introduction:** Recommendations from physicians and peers play a crucial role in promoting vaccination. This study evaluated differences in recommendations to others regarding four vaccines with varying efficacy (seasonal influenza, measles, human papillomavirus [HPV], and coronavirus disease 2019 [COVID-19]) between physicians and the general public and examined the impact of birth-year-based vaccination policy changes on these recommendations.

**Methods:** This cross-sectional study was conducted in February 2026 among 492 physicians and 5,252 members of the general public in Japan. Consistency in recommendations across the four vaccines was assessed using the intraclass correlation coefficient (ICC[3,1]), and group differences were examined using a two-way mixed-design analysis of covariance. Multilevel regression discontinuity analyses were performed to evaluate the effects of birth-year-based vaccination policy. *Results:* Physicians showed significantly stronger recommendations to others than the general public, and their recommendation patterns generally reflected vaccine efficacy. However, physicians showed lower consistency across vaccine types than the general public (ICC[3,1]), driven primarily by heterogeneity in COVID-19 vaccine recommendations. Regression discontinuity analyses showed that birth-year-based vaccination policy, including routine vaccination opportunities, was significantly associated with recommendations to others for measles and HPV vaccines, independently of perceived benefits and risks.

**Conclusion:** To improve vaccination coverage from a public health perspective, it is important for physicians to provide effective vaccination recommendations to the general public on a broader scale; however, it is also necessary to address the heterogeneity in vaccine-specific recommendation patterns among physicians, as observed for COVID-19. Routine vaccination opportunities may increase vaccination coverage not only through the routine vaccination program itself but also through peer effects among the general public. Vaccination policy may therefore influence vaccination coverage not only in the current generation but also in future generations. Designing vaccination policy should consider its long-term impact on future vaccination coverage as well as herd immunity.

## 1. Introduction

Vaccination is an essential countermeasure against infectious diseases. Vaccination benefits both vaccinated individuals and society. At the individual level, vaccines prevent infectious diseases, and some vaccines, such as those against measles and human papillomavirus (HPV), can prevent infection [1–3]. At the societal level, vaccination reduces the burden on healthcare systems by decreasing the number of infected individuals and severe cases [ms4]. Vaccination also suppresses outbreaks of infectious diseases through herd immunity. However, to fully realize these benefits, obstacles such as vaccine hesitancy must be overcome [5].

One way to address these barriers is through vaccine recommendations from others. Vaccination behavior is influenced not only by benefits and costs but also by others’ recommendations. Vaccine recommendations from physicians encourage vaccination among the general public [6]. Conversely, when physician recommendations are absent, vaccination uptake decreases among the general public [7]. Peer effects have also been observed among the general public, whereby one person’s vaccination encourages vaccination among others, possibly in part through interpersonal information sharing [8, 9]. As part of this interpersonal information sharing, vaccine recommendations from the general public may also encourage others’ vaccination behavior.

Vaccine recommendations from others may promote vaccination behavior in future generations. When parents are vaccinated, they are more likely to vaccinate their children with the same vaccine [10]. Therefore, when others’ recommendations encourage parents to be vaccinated, intentions to vaccinate children may also increase. In addition, vaccine recommendations from physicians directly increase parents’ intentions to vaccinate their children [11]. Because stronger intentions may increase vaccination uptake, vaccine recommendations from others may bring benefits to individuals and society in future generations.

It is therefore important to understand how recommendations to others are formed. This study focuses on institutional factors, such as routine immunization programs, that shape them. This focus has two implications. First, it fills a gap in the existing literature. Prior research has largely focused on intrapersonal factors such as perceptions of vaccine benefits and risks [10, 12, 13]. By contrast, vaccination policy may influence the formation of recommendations to others through the provision of vaccination opportunities and government-endorsed social norms. Second, it has policy implications. Vaccination policy has typically been evaluated based on how they affect uptake in the target generation. However, if institutional factors shape recommendations to others, policy evaluation should also consider that policy may indirectly affect vaccination behavior in future generations through changes in recommendations to others, rather than only uptake in the current generation.

This study had three aims, focusing on four vaccines in Japan (seasonal influenza, measles, HPV, and coronavirus disease 2019 [COVID-19]). These vaccines were selected because their efficacy and vaccination programs differ from one another. First, we verified the validity of the questionnaire items regarding recommendations to others among physicians and the general public by examining their relationship with other items, such as agreement with vaccination and vaccination history. Second, we assessed the consistency of recommendations to others separately among physicians and the general public and compared recommendations to others between physicians and the general public and across the four vaccines, along with the associated factors, namely, perceived benefits and risks of the vaccines. Third, we examined the impact of vaccination policy on recommendations to others and whether this impact differed between physicians and the general public. Physicians are likely to have greater scientific knowledge about vaccines than the general public. Therefore, the same vaccination policy may not affect physicians and the general public in the same way.

To address the third aim, we used an econometric method known as regression discontinuity analysis. This analysis is a quasi-experimental method that estimates policy effects by exploiting discontinuities at policy eligibility thresholds and is applicable in public health [14, 15]. In this study, we exploited birth-year-based differences in vaccination policy to estimate their effects on recommendations to others.

## 2. Methods

### 2.1. Ethics

This study was approved by the Ethics Committee of the Center for Infectious Disease Education and Research at the University of Osaka (approval numbers: 2025CRER0120; 2026CRER0415-2). All participants provided informed consent prior to participation and received points redeemable for merchandise.

### 2.2. Participants

This study was conducted using a cross-sectional design. Details of this survey are described in a previous study [16]. Briefly, online surveys were conducted among physicians from February 17 to 18, 2026, and among members of the general public from February 17 to 25, 2026.

All participants were registered panel members of Nikkei Research Inc. In Japan, two out of three physicians are registered with Nikkei Research’s physician panel. Their status as physicians was verified through a rigorous physician authentication system, which includes checking graduation records and sending mail to their places of employment.

The target sample size was 440 physicians and 5,000 members of the general public (625 respondents from each of eight regions), exceeding the required sample size (i.e., 384 respondents) [17]. For the general public, responses were collected by region to match the age and gender distribution of the whole Japanese population. Of the 1,041 physicians and 6,794 members of the general public who initially responded to the survey, exclusions were made based on consent, age, gender, dropouts during the survey, and quota being filled, resulting in a final sample of 492 physicians and 5,252 members of the general public (Figure 1). During the survey, two directed question scale (DQS) items were administered to identify inattentive respondents [18, 19]. Respondents who failed either DQS, the general public reporting their occupation as “physician,” and physicians reporting an age younger than 24 years were classified as “inattentive or ineligible respondents.” There were 101 (20.5%) inattentive or ineligible respondents among physicians and 2,017 (38.4%) among the general public.

**Figure 1.**
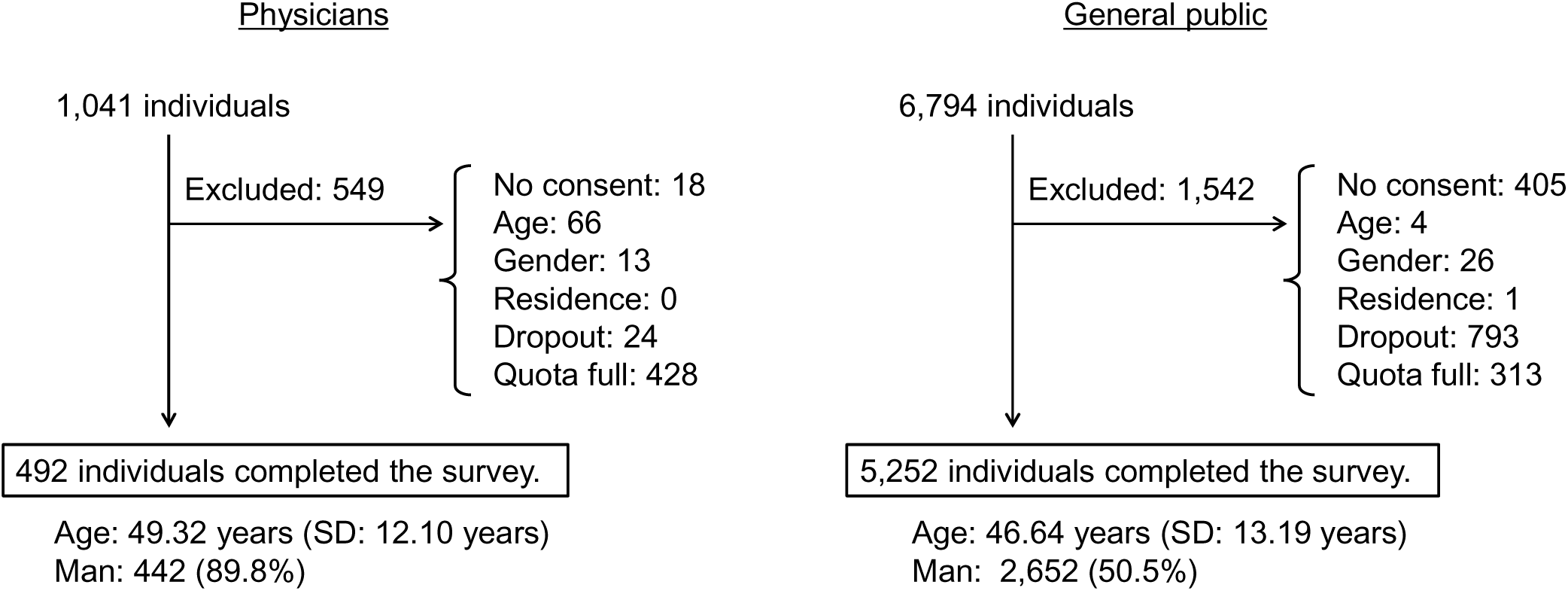
Participant flow diagram. SD: standard deviation.

### 2.3. Vaccines

This study analyzed four types of vaccines: seasonal influenza, measles, HPV, and COVID-19. The World Health Organization assessments of these vaccines [20–23] have also been summarized previously [16]. Briefly, measles and HPV vaccines are highly effective, seasonal influenza vaccines provide moderate protection that varies by season, and COVID-19 vaccines remain highly effective in preventing severe disease despite reduced effectiveness against infection after the emergence of the Omicron variant.

In Japan, vaccination is based on individual choice, although some vaccines are included in the routine immunization program under the Immunization Act. Historical system changes vary by vaccine. For seasonal influenza, school-based vaccination began in 1962, and was designated a temporary vaccination under the Act in 1976 [24]. The vaccination system was revised in 1987, and removed from the Act in 1994, and school-based mass vaccination was abolished. Currently, annual voluntary vaccination with one dose is recommended for adults.

For measles, routine vaccinations under the Act began in 1978, but after the 1994 amendment to the Act, the program shifted to individual vaccinations. Since 2006, two routine doses of the MR vaccine (measles-rubella combination vaccine) have been administered to children. As a special measure, individuals born between April 2, 1990 and April 1, 2000 were eligible for a second dose for a five-year period beginning April 1, 2008. Consequently, many people—particularly those born on or before April 1, 1990—received no or only one dose and therefore lack sufficient antibodies.

In fiscal year 2010, HPV vaccination was offered to girls in the first year of junior high school to the first year of high school. In April 2013, active recommendation began for girls in their sixth grade of elementary school through their first year of high school; however, this active recommendation was suspended in June 2013. Active recommendation resumed in April 2022, and currently, a two- or three-dose schedule is recommended. For girls born between April 2, 1997 and April 1, 2009, catch-up vaccinations were conducted from April 2022 through March 2026 (with the first dose administered by March 2025).

COVID-19 vaccination began in February 2021 free of charge. In April 2024, the program transitioned to annual routine vaccination for individuals aged 65 and older, as well as those aged 60–64 with underlying medical conditions who are at high risk of severe illness. For all other individuals, the vaccination is at their own expense.

### 2.4. Survey items

Participants rated their recommendation of each vaccine to others, including family, friends, and colleagues, on an 11-point Likert scale (0 = not recommend at all to 10 = strongly recommend); this outcome was hereafter referred to as recommendations to others. To assess its validity, participants also reported their agreement with each vaccination (0 = strongly disagree to 10 = strongly agree) and their vaccination history (for seasonal influenza, within the past two years; for other vaccines, lifetime experience; response options, yes, no, or don’t know).

Independent variables were age, participant type (physician or the general public), perceived benefits of vaccination (perceived effectiveness in preventing infection and severe disease), and perceived risks (perceived likelihood of mild and severe adverse events). Perceived benefits and risks were assessed using two items on an 11-point Likert scale. The covariates were gender, having children, and having a partner. These variables were selected based on previous studies demonstrating associations with vaccination intention, vaccination history, and recommendations to others [10, 25–27].

### 2.5. Statistical analysis

First, Spearman-Brown coefficients were calculated for perceived benefits and risks for each vaccine and confirmed to be acceptable (benefits: seasonal influenza, 0.82; measles, 0.89; HPV, 0.88; COVID-19, 0.86; risks: seasonal influenza, 0.66; measles, 0.71; HPV, 0.72; COVID-19, 0.59); therefore, the mean values were used in subsequent analyses.

Second, to assess the validity of recommendations to others, their associations with vaccination agreement and vaccination history were examined separately by participant type and vaccine. For vaccination agreement, we used Spearman’s correlation; for vaccination history, we performed a one-way analysis of variance and calculated the effect sizes R² and η².

Third, intraclass correlation coefficients [ICC](3,1) were calculated using a two-way mixed-effects model with vaccine type as a fixed effect to assess the consistency of individual recommendations to others across vaccines. The analysis was repeated after excluding each vaccine in turn.

Fourth, differences in perceived benefits, perceived risks, and recommendations to others between physicians and the general public and across vaccines were analyzed using a two-way mixed-design analysis of covariance (between subjects: participant type; within subjects: vaccine type) with age and gender as covariates. Because Mauchly’s test of sphericity was significant (*P* < 0.001), Greenhouse–Geisser corrections were applied. Simple main effects were adjusted using the Bonferroni correction.

Fifth, to examine the effect of the vaccination policy on recommendations to others, we conducted regression discontinuity analyses, which estimate causal effects of policy changes by exploiting differences in eligibility around policy thresholds [28]. We focused on birth-year-based differences in the vaccination eligibility. As mentioned above, for the measles vaccine, individuals born on or after April 2, 1990 (aged ≤ 35 in February 2026, when the survey was conducted) were offered two doses. Furthermore, measles and seasonal influenza vaccination shifted from routine to individual vaccination following the 1994 amendment to the Act. The cohort entering elementary school in 1994 consisted of individuals born between April 2, 1987 and April 1, 1988. However, even after the transition to individual vaccination, social norms regarding mass vaccination may have persisted for a certain period. Therefore, in the regression discontinuity analysis, the participants were divided into two main categories: those aged 35 or younger and those aged 36 or older. Furthermore, we excluded those aged 34 to 37 and created additional categories for those aged 20 to 33 and those aged 38 and older. Additionally, regarding the HPV vaccine, since those who were first-year high school students in April 2010—when vaccination opportunities began—correspond to those born between April 2, 1994 and April 1, 1995, we also divided the sample into those aged 31 and younger and those aged 32 and older.

Specifically, the following analyses were conducted. First, as a preliminary step, regression discontinuity analyses were performed for each of the four vaccines, with recommendations to others as the outcome variable. The independent variables were age (centered at 36 years), pre-policy status (1 = 36 years or older, 0 = 35 years or younger), age × pre-policy status, physician (ref: general public), physician × age, physician × pre-policy status, perceived benefits, and perceived risks. Gender, having children, and having a partner were also included as covariates. While the regression coefficients of the policy status-related terms (age, pre-policy status, age × pre-policy status) showed different patterns across vaccines, none of the three physician-related interaction terms was significant for any vaccine. Therefore, multilevel regression analysis was subsequently performed. The independent variables in the multilevel regression analysis were age (centered at 36 years), pre-policy status (1 = 36 years or older, 0 = 35 years or younger), age × pre-policy status, physician, each vaccine (ref: measles), vaccine × age, vaccine × pre-policy status, vaccine × age × pre-policy status, perceived benefits, and perceived risks; the covariates were gender, having children, and having a partner. Individual IDs were included as random effects. As primary analyses, we conducted multilevel regression analyses among all participants and among those excluding participants aged 34–37. The models adjusting for perceived benefits and risks estimate the effect of the vaccination policy that is not explained by these perceptions (i.e., the direct effect), whereas the models without these covariates estimate the total effect. We therefore conducted both analyses and compared the results to assess the extent to which the policy effect was mediated by perceived benefits and risks. As a sensitivity analysis, we performed a multilevel regression analysis on participants excluding inattentive or ineligible respondents. As a second sensitivity analysis, we also conducted a standard multiple regression analysis specifically for the HPV vaccine, using a dichotomous classification of participants aged 31 or younger versus 32 or older. The independent variables were age (centered at 32 years), pre-policy status (1 = 32 years or older, 0 = 31 years or younger), age × pre-policy status, physician, perceived benefits, and perceived risks; the covariates were gender, having children, and having a partner. Based on the obtained regression coefficients, we calculated the age-related slope of recommendations to others among those aged 35 or younger and those aged 36 or older, as well as the difference in recommendations to others associated with the policy status. The 95% confidence interval (CI) was calculated using the delta method.

Variance inflation factors were estimated to confirm low multicollinearity. All analyses were performed using IBM SPSS version 28 and R version 4.5.3.

## 3. Results

### 3.1. Validity of Items regarding Recommendations to Others

Recommendations to others reflect respondents’ own attitudes and behavior. Table 1 presents associations between recommendations to others and agreement with vaccination by vaccine type and participant type (physicians or the general public), showing strong positive correlations between the two measures (physicians, 0.782–0.861; general public, 0.663–0.726). Those with a vaccination history had higher recommendations to others than those without (Table S1). In particular, seasonal influenza showed the highest effect size η^2^ (physicians, 0.149; general public, 0.135), followed by measles (0.078; 0.076), COVID-19 (0.039; 0.050), and HPV (0.013; 0.013).

**Table 1.**
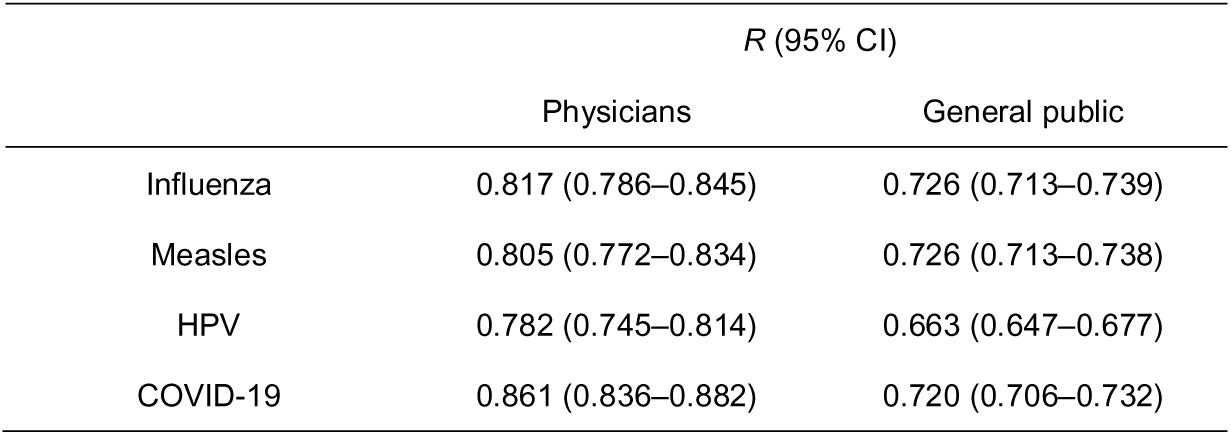
Correlations between recommendations to others and vaccination agreement. CI: confidence interval.

### 3.2. Differences in Recommendations to Others between Physicians and the General Public

Based on ICC(3,1), physicians showed lower consistency in recommendations to others across the four vaccine types than the general public (Table 2; physicians, 0.465; general public, 0.719). However, when COVID-19 was excluded, the ICC(3,1) increased to 0.616 among physicians. Physicians also perceived greater benefits and risks of vaccination and recommended vaccination to others more strongly than the general public, irrespective of vaccine type (Table 3). Among physicians, the highest recommendations to others were measles (0.866), followed by HPV (0.813) and seasonal influenza (0.795), and finally COVID-19 (0.596). In contrast, among the general public, the recommendations to others were the highest for measles (0.543), followed by seasonal influenza (0.521), HPV (0.507), and COVID-19 (0.425). For both physicians and the general public, perceived benefits generally showed patterns similar to those of recommendations to others, whereas perceived risks showed the opposite pattern.

**Table 2.**
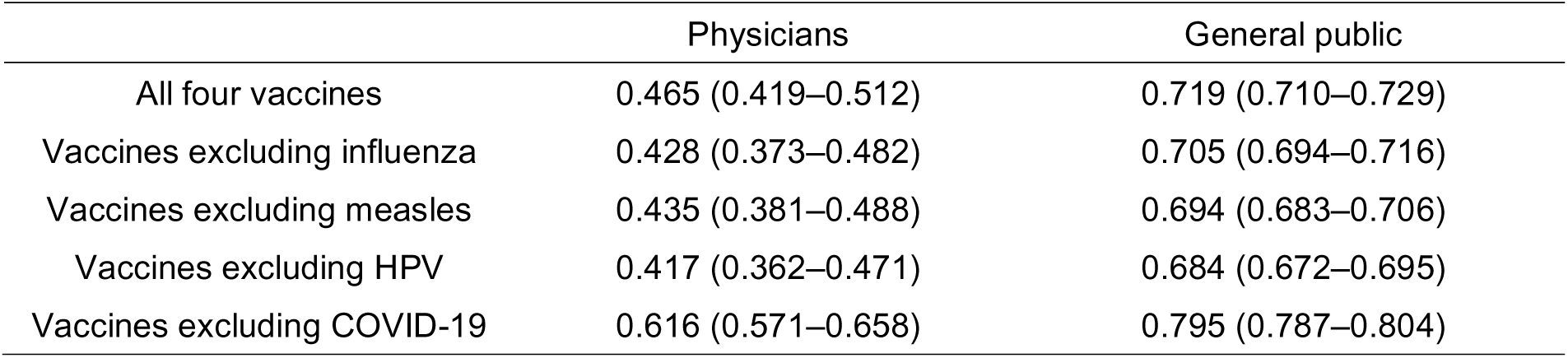
Relative consistency of recommendations to others within individuals across the vaccine types (ICC(3,1)). Values in parentheses indicate 95% confidence intervals.

**Table 3.**
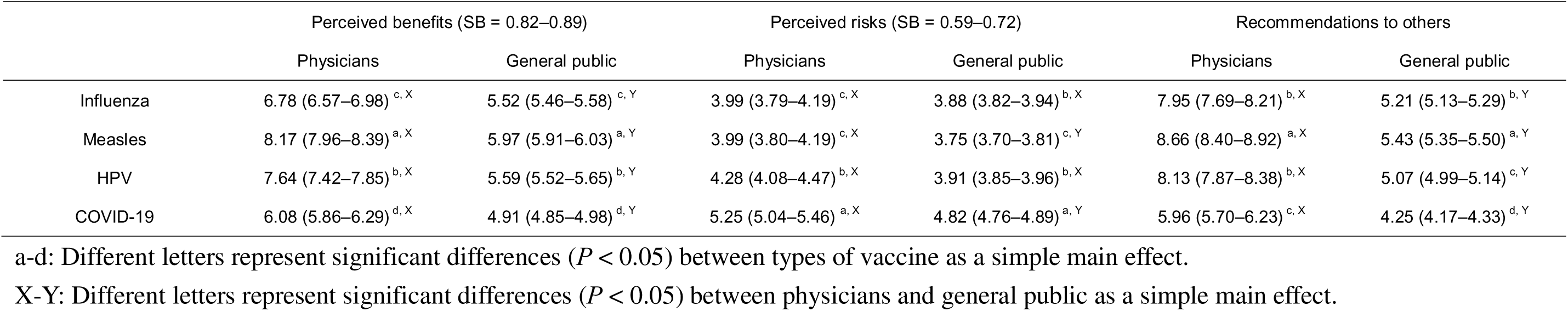
Perceived benefits and risks of vaccination, and recommendations to others. Data are presented as means (95% confidence intervals). Simple main effects are adjusted by Bonferroni correction. Means were adjusted for age (46.87 years) and gender (male = 0.54). Perceived benefits: interaction, *P* < 0.001, partial η^2^ = 0.016; perceived risks: interaction, *P* = 0.001, partial ^2^ = 0.001; recommendation to others: interaction, *P* < 0.001, partial η^2^ = 0.014. SB: Spearman Brown coefficient.

|  | Perceived benefits (SB = 0.82–0.89) |  | Perceived risks (SB = 0.59–0.72) |  | Recommendations to others |  |
| --- | --- | --- | --- | --- | --- | --- |
|  | Physicians | General public | Physicians | General public | Physicians | General public |
| Influenza | 6.78 (6.57–6.98)<sup>c, X</sup> | 5.52 (5.46–5.58)<sup>c, Y</sup> | 3.99 (3.79–4.19)<sup>c, X</sup> | 3.88 (3.82–3.94)<sup>b, X</sup> | 7.95 (7.69–8.21)<sup>b, X</sup> | 5.21 (5.13–5.29)<sup>b, Y</sup> |
| Measles | 8.17 (7.96–8.39)<sup>a, X</sup> | 5.97 (5.91–6.03)<sup>a, Y</sup> | 3.99 (3.80–4.19)<sup>c, X</sup> | 3.75 (3.70–3.81)<sup>c, Y</sup> | 8.66 (8.40–8.92)<sup>a, X</sup> | 5.43 (5.35–5.50)<sup>a, Y</sup> |
| HPV | 7.64 (7.42–7.85)<sup>b, X</sup> | 5.59 (5.52–5.65)<sup>b, Y</sup> | 4.28 (4.08–4.47)<sup>b, X</sup> | 3.91 (3.85–3.96)<sup>b, X</sup> | 8.13 (7.87–8.38)<sup>b, X</sup> | 5.07 (4.99–5.14)<sup>c, Y</sup> |
| COVID-19 | 6.08 (5.86–6.29)<sup>d, X</sup> | 4.91 (4.85–4.98)<sup>d, Y</sup> | 5.25 (5.04–5.46)<sup>a, X</sup> | 4.82 (4.76–4.89)<sup>a, Y</sup> | 5.96 (5.70–6.23)<sup>c, X</sup> | 4.25 (4.17–4.33)<sup>d, Y</sup> |

a-d: Different letters represent significant differences ( $P < 0.05$ ) between types of vaccine as a simple main effect.
X-Y: Different letters represent significant differences ( $P < 0.05$ ) between physicians and general public as a simple main effect.

### 3.3. Impact of Vaccination Policy on Recommendations to Others

Figure 2 shows age profiles of recommendations to others by vaccine type and participant type. Apparent discontinuities at the age thresholds corresponding to vaccination policy changes were observed for some vaccine types, suggesting possible policy effects.

**Figure 2.**
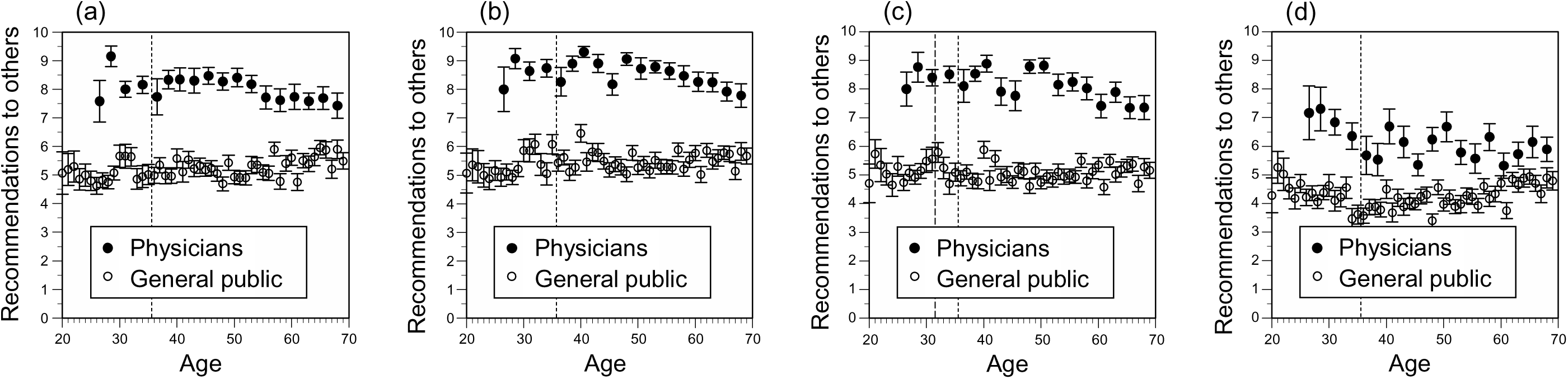
Associations between recommendations to others and age. (a) Influenza, (b) measles, (c) HPV, (d) COVID-19. Error bars represent standard errors. The dashed line indicates the age threshold for the vaccination policy change. Physicians’ ages were grouped into 2- to 3-year intervals.

We formally tested this hypothesis using regression discontinuity analyses. First, as a preliminary step, we estimated regression discontinuity analyses for each vaccine type (Table S2). The regression coefficients of the physician-related interaction terms (physician × age, physician × pre-policy status, physician × age × pre-policy status) were not significant for any vaccine (*P* > 0.05), whereas the regression coefficients of the policy status-related terms (age, pre-policy status, age × pre-policy status) showed different patterns across vaccines.

We then performed the multilevel regression analysis (Tables 4 and S3). Because heterogeneity in discontinuities across vaccine types was accounted for, the coefficient on pre-policy status represents the impact of the vaccination policy change on the recommendations to others for measles. Using the full sample, the estimated coefficient on pre-policy status was −0.28 (*P* = 0.066). The analysis excluding participants aged 34–37 years strengthened this association (coefficient = −0.40, *P* = 0.048) whereas in the analysis without adjusting for perceived benefits and risks, the coefficient remained nearly identical but was no longer statistically significant (coefficient = −0.31, *P* = 0.110). Furthermore, although this finding was not robust, the effect of vaccination policy change differed across vaccine types. In particular, across vaccine types, the effects for seasonal influenza and COVID-19 shifted in a positive direction relative to measles.

Using linear combination tests of the estimates from Tables 4 and S2, Table 5 presents these patterns of heterogeneity more clearly. Based on the analysis excluding participants aged 34–37 years, the coefficients of pre-policy status were negative and statistically significant at the 5% level for measles and HPV (−0.40 [−0.13] for measles and −0.45 [−0.15] for HPV, where values in brackets represent unstandardized coefficients divided by the standard deviation of the outcome). The analysis without adjustment for perceived benefits and risks showed nearly identical coefficients but wider confidence intervals than the adjusted analysis. Based on the vaccine-stratified regression discontinuity analyses, this effect was concentrated among the general public. In particular, the vaccination policy change was associated with recommendations to others for the measles vaccine among the general public (for measles: physicians −0.01 [−0.01] vs. general public −0.34 [−0.12]). Furthermore, among physicians, a significant negative association was observed between age and recommendations to others for seasonal influenza for those aged 36 and older. Among the general public, a significant negative association was observed between age and recommendations to others for COVID-19 for those aged 35 and younger, while a positive association—though at the borderline level of significance—was observed for those aged 36 and older.

**Table 4.**
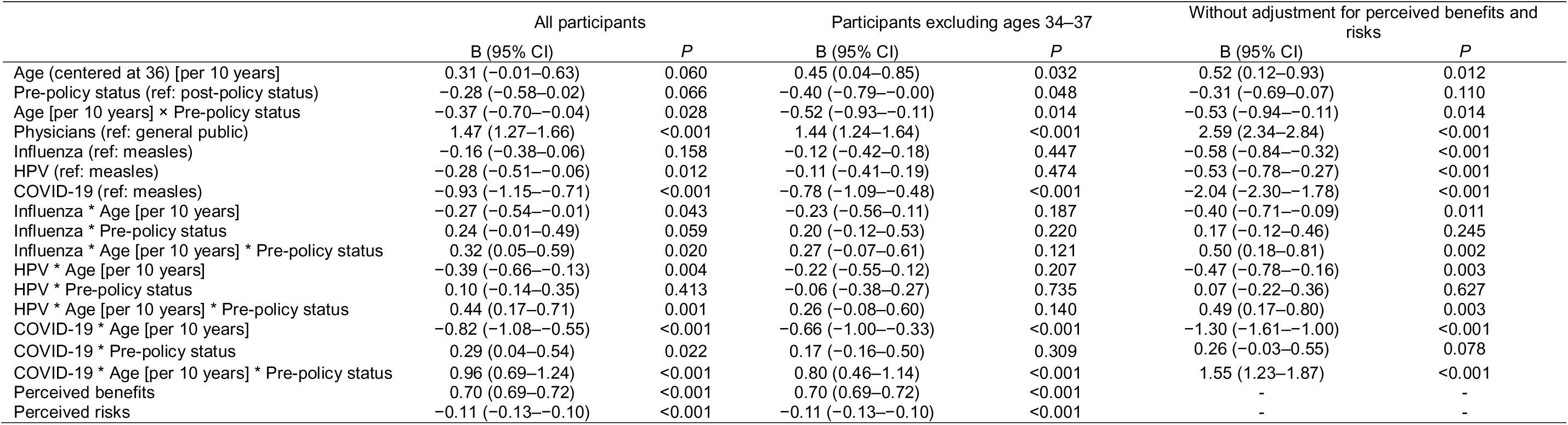
Unstandardized regression coefficients for recommendations to others. Random effects are as follows: all participants, between-person standard deviation (σ_u_) = 1.84, residual standard deviation (σ_e_) = 1.34, intraclass correlation coefficient (ICC) = 0.65; participants excluding ages 34–37, σ_u_ = 1.83, σ_e_ = 1.33, ICC = 0.65; without adjustment for perceived benefits and risks, σ_u_ = 2.41, σ_e_ = 1.56, ICC = 0.71. Variance inflation factors for covariates, excluding variables used in the regression discontinuity design (i.e., age, pre-policy status, vaccine types, and all associated interaction terms): ≤ 1.52 for all participants; ≤ 1.51 for participants excluding ages 34–37; ≤ 1.52 for without adjustment for perceived benefits and risks. CI: confidence interval. Full results with the intercept and covariates, including gender, children, and partner, are shown in Table S3.

**Table 5.**
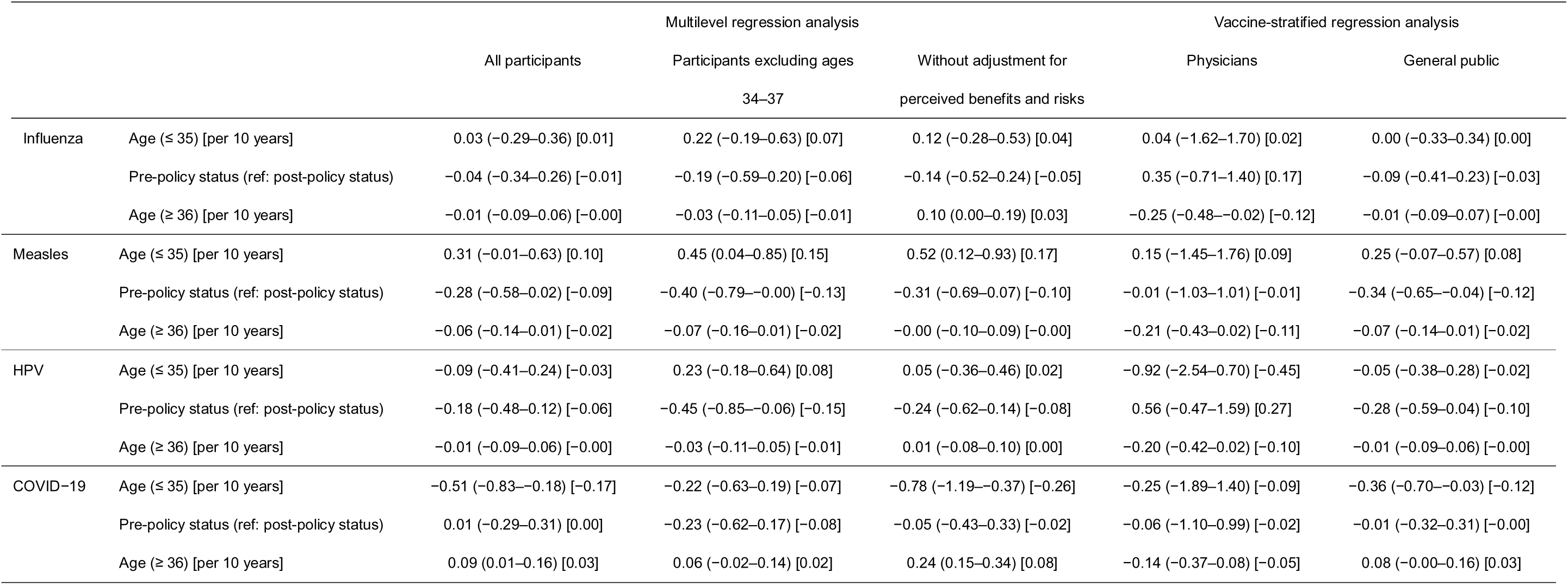
Unstandardized coefficients (linear combinations) for recommendations to others. Values in parentheses represent 95% confidence intervals derived using the delta method. Values in brackets represent unstandardized coefficients divided by the standard deviation of the corresponding outcome distribution in each analysis population.

These findings were further supported by additional analyses (Tables S4 and S5).

## 4. Discussion

This study assessed recommendations to others for four vaccines among physicians and the general public. Differences in recommendation patterns were observed between groups and across vaccines, and policy-related differences were identified for measles and HPV vaccines among the general public.

Recommendations to others showed a high correlation with vaccination agreement for both physicians and the general public (physicians: 0.782–0.861; general public: 0.663–0.726), confirming their validity. Regarding the association with vaccination history, for both physicians and the general public, the effect size η² was highest for seasonal influenza, followed by measles, COVID-19, and HPV. Participants with a history of seasonal influenza vaccination reported higher recommendations to others, and the effect size η² was also high (physicians: 0.149; general public: 0.135). For measles and HPV, the low effect size can be explained by the fact that the target population included many people with no vaccination history due to the vaccination policy. For the COVID-19 vaccine, since the recommended target population differed before and after the Omicron variant [23], the association between vaccination history and current recommendations to others for COVID-19 vaccination was also reasonably low.

The ICC(3,1) was high among the general public (0.719) but lower among physicians (0.465), indicating that recommendation patterns were less consistent across vaccines among physicians. Among the general public, recommendations appeared to move largely along a single pro- versus anti-vaccination dimension, yielding high consistency, whereas physicians evaluated each vaccine more independently, which in itself reduces consistency. This finding is consistent with previous studies reporting differences in vaccination attitudes among physicians [27, 29]. Importantly, this lower consistency was concentrated in COVID-19: excluding it produced a markedly larger increase in ICC(3,1) among physicians (0.465 to 0.616; +0.151) than among the general public (0.719 to 0.795; +0.076). This asymmetry indicates that the substantially lower consistency among physicians was driven primarily by between-physician heterogeneity in the evaluation of the COVID-19 vaccine.

Although recommendation patterns differed among individuals, overall, among physicians, the recommendations to others were highest for measles, followed by HPV and seasonal influenza, with COVID-19 having the lowest score. Among the general public, however, the highest recommendations to others were for measles, followed by seasonal influenza, HPV, and COVID-19. This ranking generally corresponded to high perceived benefits and low perceived risks; both physicians and the general public perceived measles as having the highest perceived benefits and lowest perceived risks, while COVID-19 was perceived as the opposite. This ranking is broadly consistent with the WHO assessments of vaccine efficacy [20–23]. The general public tended to be slightly less likely to recommend the HPV vaccine; one possible reason for this is the suspension of active recommendation for HPV vaccination in Japan due to adverse events (from June 2013 to March 2022) [30]. Regression analysis revealed that recommendations to others were positively associated with perceived benefits and negatively associated with perceived risks, which was consistent with findings from previous studies on vaccination intentions, history, and recommendations to others among both physicians and the general public [10, 25–27]. Interestingly, regardless of the vaccine type, physicians reported higher scores than the general public not only for recommendations to others and perceived benefits but also for perceived risks. These results indicate that physicians recommend vaccination to others while recognizing potential risks. One possible reason for this is that, as professionals, physicians are generally more conscious of and knowledgeable about both the benefits and risks of vaccination; however, as mentioned earlier, it is important to note that substantial heterogeneity exists in vaccine-specific recommendation patterns among physicians.

Regression discontinuity analysis showed that policy changes were associated with increases in recommendations to others (negative regression coefficients for pre-policy status) for the measles and HPV vaccines. This difference was particularly pronounced in the analysis of the participants excluding ages 34–37 for both vaccines, and in the sensitivity analysis for the HPV vaccine that distinguished between those aged 31 and younger and those aged 32 and older. The policy effect persisted after adjustment for perceived benefits and risks, and the total-effect estimates were similar to the direct-effect estimates (measles: −0.31 vs. −0.28), indicating little mediation through these perceptions. Although the coefficients were not statistically significant in the unadjusted analysis, this was likely due to wider confidence intervals rather than differences in the estimated effects. This suggests that perceived benefits and risks improved the precision of the estimates by explaining residual variance rather than altering the estimated policy effect. Because prior research on recommendations to others has focused predominantly on such intrapersonal perceptions [10, 12, 13], these findings suggest that vaccination policy operates through additional pathways, such as social norms associated with inclusion in the routine immunization program. Furthermore, when analyzing the data by physician and general public, the impact of the policy change on recommendations to others for measles vaccination was significant only among the general public.

The fact that the analysis excluding those aged 34–37 showed a more pronounced effect suggests that social norms may persist even after policy changes. Regarding the measles vaccine, individuals born after the policy change were provided with two opportunities for routine vaccination (including special measures). For the HPV vaccine as well, catch-up vaccinations were administered in addition to the vaccination opportunities available since 2010. Therefore, the provision of routine vaccination opportunities may have contributed to higher recommendations to others. In contrast, no differences in recommendations to others between pre- and post-policy status were observed for seasonal influenza and COVID-19. For COVID-19, this result is attributable to the fact that there were no birth-year-based policy changes for the age groups covered in this study. For seasonal influenza, the impact of policy changes based on birth year was likely small because annual or biannual vaccination is currently recommended.

Among physicians, no increase in recommendations to others for measles and HPV vaccination was observed following policy changes; although not statistically significant, a trend toward the opposite direction compared to the general public was observed (regression coefficients of pre-policy status for measles: physicians −0.01 vs. general public −0.34; for HPV: physicians 0.56 vs. general public −0.28). One possible interpretation is that their professional knowledge regarding vaccination may have attenuated the impact of policy changes based on birth year; however, this requires verification through analysis using greater statistical power.

Furthermore, among the general public aged 35 and younger, the recommendations to others for COVID-19 vaccination decreased significantly with increasing age, whereas among those aged 36 and older, a trend toward an increase with age (borderline significance) was observed. Since previous studies—including those after the Omicron variant emerged—have generally indicated that older individuals have higher COVID-19 vaccination intentions and uptake [25, 31, 32], the positive association between age and recommendations to others observed in this study for those aged 36 and older is consistent with these findings. In contrast, focusing on the under-35 age group, a 2021 U.S. study targeting individuals aged 18 to 39 found that the proportion of respondents who answered “probably or definitely will not get vaccinated” tended to be lower among younger participants [33]. Regarding intentions to receive COVID-19 booster shots in Japan, there was no significant difference between those in their 20s and those in their 30s [34, 35]. Furthermore, in Japan, young people are more likely to view altruistic considerations rather than their own interests as the main driving force behind their willingness to get vaccinated against COVID-19 [36]. Therefore, regardless of their own vaccination intentions, among those aged 35 and under, younger people may have been more likely to recommend vaccination to others.

Among physicians, the recommendations to others for seasonal influenza vaccination decreased significantly among those aged 36 and older. In previous studies on the 2009 influenza A/H1N1 pandemic and seasonal influenza, the association between physicians’ age and their recommendations to others was not clear [12, 13]. A review article on healthcare providers’ vaccine recommendation behavior reported that no consistent results have been obtained regarding associations with age or years of medical experience [27]. This study suggested that older physicians may have been more hesitant to recommend vaccination to others; however, caution is warranted regarding the generalizability of these findings.

This study highlighted several social implications. First, physicians were more likely than the general public to recommend vaccination to others, and they were more likely to recommend vaccines with higher efficacy. Since the public’s intention to get vaccinated is associated with information availability and recommendations from physicians [6, 25], it is important for public health that physicians recommend highly effective vaccines. However, as observed for COVID-19, the heterogeneity in individual physicians’ vaccine-specific recommendations to others could pose a challenge for communication strategies aimed at encouraging a wider range of the public to get vaccinated. It is important not only to promote the sharing of evidence regarding vaccines among physicians but also to recognize that the public may receive different messages depending on the physician providing the recommendation. Second, this study suggests that the vaccination policy influences the public’s willingness to recommend vaccination to others. In particular, it is important to note that the provision of routine vaccinations and catch-up vaccination opportunities may positively contribute to recommendations to others. The routine vaccination program is likely to increase vaccination coverage not only through its direct effect on vaccination uptake but also indirectly through peer effects among the general public. Furthermore, given that the general public’s recommendations to others are a key factor in vaccination intentions among family members, especially children [37], vaccination policy has the potential to influence vaccination rates not only for the current generation but also for future generations. It is important to consider the design of vaccination policy not only in terms of achieving herd immunity at a given point in time but also from the perspective of its impact on future generations’ vaccination coverage.

This study has several limitations. First, the use of survey company panels may have introduced selection bias. Although physician panel registration was relatively controlled and the general public panel was recruited to match Japan’s age and gender distribution, biases related to other characteristics may remain. Second, because the study was conducted in Japan, the generalizability of the findings to other regions should be interpreted with caution. Third, although regression discontinuity analysis was applied, the cross-sectional design limits causal interpretation.

## 5. Conclusions

This study evaluated recommendations to others regarding four vaccines with varying efficacy among physicians and the general public. While physicians recommended vaccination to others more strongly than the general public, and the strength of their recommendations was consistent with vaccine efficacy, substantial heterogeneity in vaccine-specific recommendation patterns among individual physicians was identified, particularly for COVID-19. To improve vaccination coverage for public health purposes, it is important for physicians to provide stronger recommendations for effective vaccines to the general public; however, it is also necessary to consider and address the heterogeneity in vaccine-specific recommendation patterns among physicians. Furthermore, regression discontinuity analysis suggested that vaccination policy changes, including the timing of routine vaccination opportunities based on birth year, influence recommendations to others for measles and HPV vaccination. Vaccination policy might influence vaccination coverage not only through direct program effects but also indirectly through peer effects, with potential impacts extending to future generations, including children. Therefore, it is important to design vaccination policy not only with regard to achieving herd immunity at a given point in time but also from the perspective of its potential impacts on future generations’ vaccination coverage.

## Data availability

Data will be made available upon request.

## AI Statement

The authors used DeepL, ChatGPT, and Claude to improve the English language of this manuscript. The manuscript was initially drafted by the authors, who critically reviewed and revised all AI-assisted edits. The authors take full responsibility for the content of this publication.

## CRediT authorship statement

Conceptualization: Michio Murakami and Fumio Ohtake

Methodology: Michio Murakami and Fumio Ohtake

Formal analysis: Michio Murakami and Hiroki Kato

Investigation: Michio Murakami and Fumio Ohtake

Data curation: Michio Murakami

Writing - Original Draft: Michio Murakami and Hiroki Kato

Writing - Review & Editing: Michio Murakami, Hiroki Kato and Fumio Ohtake

Visualization: Michio Murakami

Project administration: Fumio Ohtake

Funding acquisition: Michio Murakami, Hiroki Kato, and Fumio Ohtake

