## Supplementary Materials for "Vaccination recommendations to others among physicians and the general public: effects of birth-year-based vaccination policy changes assessed by regression discontinuity analysis"

Table S1. Associations between recommendations to others and vaccination history. CI: confidence interval.

| | Physicians | | | | General public | | | |
|---|---|---|---|---|---|---|---|---|
| | Mean (95%CI) [n] | | | $\eta^2$ (95% CI) | Mean (95%CI) [n] | | | $\eta^2$ (95% CI) |
| | Yes | No | Don't' know | | Yes | No | Don't' know | |
| Influenza | 8.23 (8.06–8.39) [452] | 5.30 (4.35–6.25) [37] | 6.33 (2.54–10.13) [3] | 0.149 (0.094–0.204) | 6.59 (6.48–6.71) [2001] | 4.36 (4.26–4.46) [3103] | 4.28 (3.83–4.74) [148] | 0.135 (0.118–0.151) |
| Measles | 8.85 (8.70–8.99) [360] | 7.93 (7.50–8.35) [109] | 7.09 (5.80–8.37) [23] | 0.078 (0.037–0.125) | 6.47 (6.34–6.61) [1851] | 4.60 (4.47–4.72) [2102] | 5.31 (5.17–5.46) [1299] | 0.076 (0.063–0.090) |
| HPV | 8.61 (8.21–9.00) [56] | 8.10 (7.90–8.30) [428] | 6.88 (5.06–8.69) [8] | 0.013 (0.000–0.036) | 5.95 (5.67–6.22) [469] | 4.89 (4.80–4.98) [3894] | 5.36 (5.18–5.53) [889] | 0.013 (0.007–0.019) |
| COVID-19 | 6.21 (5.97–6.45) [459] | 4.10 (2.91–5.29) [30] | 4.00 (-4.96–12.96) [3] | 0.039 (0.011–0.075) | 4.59 (4.50–4.68) [4056] | 2.96 (2.79–3.13) [1085] | 4.03 (3.51–4.54) [111] | 0.050 (0.039–0.061) |

Table S2. Unstandardized regression coefficients for recommendations to others (vaccine stratification). Variance inflation factors for covariates, excluding variables used in the regression discontinuity design (i.e., age, pre-policy status, physician, and all associated interaction terms): ≤ 1.53 for influenza, measles, and HPV; ≤ 1.52 for COVID-19. CI: confidence interval.

| | Influenza | | Measles | | HPV | | COVID−19 | |
|---|---|---|---|---|---|---|---|---|
| | B (95% CI) | *P* | B (95% CI) | *P* | B (95% CI) | *P* | B (95% CI) | *P* |
| Age (centered at 36) [per 10 years] | 0.00 (−0.33–0.34) | 0.977 | 0.25 (−0.07–0.57) | 0.130 | −0.05 (−0.38–0.28) | 0.759 | −0.36 (−0.70–−0.03) | 0.032 |
| Pre-policy status (ref: post-policy status) | −0.09 (−0.41–0.23) | 0.576 | −0.34 (−0.65–−0.04) | 0.028 | −0.28 (−0.59–0.04) | 0.082 | −0.01 (−0.32–0.31) | 0.957 |
| Age [per 10 years] × Pre-policy status | −0.01 (−0.36–0.33) | 0.932 | −0.32 (−0.65–0.02) | 0.062 | 0.04 (−0.30–0.37) | 0.828 | 0.44 (0.10–0.79) | 0.011 |
| Physicians (ref: general public) | 1.63 (0.63–2.63) | 0.001 | 1.29 (0.33–2.26) | 0.009 | 0.99 (0.01–1.97) | 0.047 | 1.24 (0.25–2.24) | 0.014 |
| Physicians * Age [per 10 years] | 0.03 (−1.66–1.72) | 0.972 | −0.10 (−1.73–1.54) | 0.908 | −0.87 (−2.52–0.78) | 0.303 | 0.12 (−1.56–1.80) | 0.889 |
| Physicians * Pre-policy status | 0.44 (−0.66–1.54) | 0.437 | 0.33 (−0.73–1.40) | 0.541 | 0.83 (−0.24–1.91) | 0.128 | −0.05 (−1.14–1.05) | 0.932 |
| Physicians * Age [per 10 years] * Pre-policy status | −0.27 (−1.97–1.44) | 0.760 | 0.04 (−1.69–1.61) | 0.959 | 0.68 (−0.99–2.35) | 0.423 | −0.34 (−2.04–1.35) | 0.692 |
| Perceived benefits | 0.77 (0.75–0.80) | <0.001 | 0.77 (0.74–0.79) | <0.001 | 0.71 (0.69–0.74) | <0.001 | 0.77 (0.74–0.79) | <0.001 |
| Perceived risks | −0.00 (−0.03–0.02) | 0.866 | 0.01 (−0.02–0.04) | 0.456 | −0.01 (−0.04–0.01) | 0.323 | −0.07 (−0.10–−0.04) | <0.001 |
| Man (ref: woman) | 0.10 (−0.02–0.22) | 0.109 | −0.06 (−0.17–0.06) | 0.361 | 0.22 (0.10–0.33) | <0.001 | 0.29 (0.17–0.41) | <0.001 |
| With Children (ref: without children) | 0.22 (0.06–0.37) | 0.005 | 0.29 (0.14–0.44) | <0.001 | 0.19 (0.04–0.34) | 0.011 | −0.06 (−0.21–0.09) | 0.408 |
| No answer (ref: without children) | 0.13 (−0.57–0.83) | 0.720 | 0.02 (−0.66–0.70) | 0.950 | −0.11 (−0.80–0.57) | 0.745 | −0.22 (−0.92–0.48) | 0.538 |
| With partner (ref: without partner) | 0.11 (−0.04–0.26) | 0.144 | 0.21 (0.06–0.35) | 0.005 | 0.17 (0.03–0.32) | 0.017 | 0.01 (−0.14–0.16) | 0.885 |
| No answer (ref: without partner) | 0.28 (−0.33–0.89) | 0.375 | 0.34 (−0.25–0.93) | 0.258 | 0.26 (−0.33–0.86) | 0.384 | 0.25 (−0.36–0.85) | 0.427 |
| Intercept | 0.86 (0.52–1.20) | <0.001 | 1.03 (0.70–1.37) | <0.001 | 1.10 (0.77–1.43) | <0.001 | 0.51 (0.17–0.84) | 0.003 |

Table S3. Unstandardized regression coefficients for recommendations to others. Random effects are as follows: all participants, between-person standard deviation ($\sigma_u$) = 1.84, residual standard deviation ($\sigma_e$) = 1.34, intraclass correlation coefficient (ICC) = 0.65; participants excluding ages 34–37, $\sigma_u$ = 1.83, $\sigma_e$ = 1.33, ICC = 0.65; without adjustment for perceived benefits and risks, $\sigma_u$ = 2.41, $\sigma_e$ = 1.56, ICC = 0.71. Variance inflation factors for covariates, excluding variables used in the regression discontinuity design (i.e., age, pre-policy status, vaccine types, and all associated interaction terms): ≤ 1.52 for all participants; ≤ 1.51 for participants excluding ages 34–37; ≤ 1.52 for without adjustment for perceived benefits and risks. CI: confidence interval.

| | All participants | | Participants excluding ages 34–37 | | Without adjustment for perceived benefits and risks | |
|---|---|---|---|---|---|---|
| | B (95% CI) | *P* | B (95% CI) | *P* | B (95% CI) | *P* |
| Age (centered at 36) [per 10 years] | 0.31 (−0.01–0.63) | 0.060 | 0.45 (0.04–0.85) | 0.032 | 0.52 (0.12–0.93) | 0.012 |
| Pre-policy status (ref: post-policy status) | −0.28 (−0.58–0.02) | 0.066 | −0.40 (−0.79–−0.00) | 0.048 | −0.31 (−0.69–0.07) | 0.110 |
| Age [per 10 years] × Pre-policy status | −0.37 (−0.70–−0.04) | 0.028 | −0.52 (−0.93–−0.11) | 0.014 | −0.53 (−0.94–−0.11) | 0.014 |
| Physicians (ref: general public) | 1.47 (1.27–1.66) | <0.001 | 1.44 (1.24–1.64) | <0.001 | 2.59 (2.34–2.84) | <0.001 |
| Influenza (ref: measles) | −0.16 (−0.38–0.06) | 0.158 | −0.12 (−0.42–0.18) | 0.447 | −0.58 (−0.84–−0.32) | <0.001 |
| HPV (ref: measles) | −0.28 (−0.51–−0.06) | 0.012 | −0.11 (−0.41–0.19) | 0.474 | −0.53 (−0.78–−0.27) | <0.001 |
| COVID-19 (ref: measles) | −0.93 (−1.15–−0.71) | <0.001 | −0.78 (−1.09–−0.48) | <0.001 | −2.04 (−2.30–−1.78) | <0.001 |
| Influenza * Age [per 10 years] | −0.27 (−0.54–−0.01) | 0.043 | −0.23 (−0.56–0.11) | 0.187 | −0.40 (−0.71–−0.09) | 0.011 |
| Influenza * Pre-policy status | 0.24 (−0.01–0.49) | 0.059 | 0.20 (−0.12–0.53) | 0.220 | 0.17 (−0.12–0.46) | 0.245 |
| Influenza * Age [per 10 years] * Pre-policy status | 0.32 (0.05–0.59) | 0.020 | 0.27 (−0.07–0.61) | 0.121 | 0.50 (0.18–0.81) | 0.002 |
| HPV * Age [per 10 years] | −0.39 (−0.66–−0.13) | 0.004 | −0.22 (−0.55–0.12) | 0.207 | −0.47 (−0.78–−0.16) | 0.003 |
| HPV * Pre-policy status | 0.10 (−0.14–0.35) | 0.413 | −0.06 (−0.38–0.27) | 0.735 | 0.07 (−0.22–0.36) | 0.627 |
| HPV * Age [per 10 years] * Pre-policy status | 0.44 (0.17–0.71) | 0.001 | 0.26 (−0.08–0.60) | 0.140 | 0.49 (0.17–0.80) | 0.003 |
| COVID-19 * Age [per 10 years] | −0.82 (−1.08–−0.55) | <0.001 | −0.66 (−1.00–−0.33) | <0.001 | −1.30 (−1.61–−1.00) | <0.001 |
| COVID-19 * Pre-policy status | 0.29 (0.04–0.54) | 0.022 | 0.17 (−0.16–0.50) | 0.309 | 0.26 (−0.03–0.55) | 0.078 |
| COVID-19 * Age [per 10 years] * Pre-policy status | 0.96 (0.69–1.24) | <0.001 | 0.80 (0.46–1.14) | <0.001 | 1.55 (1.23–1.87) | <0.001 |
| Perceived benefits | 0.70 (0.69–0.72) | <0.001 | 0.70 (0.69–0.72) | <0.001 | - | - |
| Perceived risks | −0.11 (−0.13–−0.10) | <0.001 | −0.11 (−0.13–−0.10) | <0.001 | - | - |
| Man (ref: woman) | 0.12 (0.02–0.22) | 0.025 | 0.09 (−0.02–0.20) | 0.098 | 0.01 (−0.13–0.14) | 0.898 |
| With Children (ref: without children) | 0.16 (0.03–0.29) | 0.016 | 0.18 (0.04–0.31) | 0.009 | 0.14 (−0.03–0.31) | 0.100 |
| No answer (ref: without children) | −0.08 (−0.68–0.52) | 0.800 | 0.00 (−0.64–0.64) | 0.994 | −0.20 (−0.98–0.58) | 0.618 |
| With partner (ref: without partner) | 0.12 (−0.01–0.24) | 0.063 | 0.13 (0.00–0.26) | 0.048 | 0.27 (0.10–0.43) | 0.001 |
| No answer (ref: without partner) | 0.23 (−0.29–0.75) | 0.384 | 0.31 (−0.24–0.86) | 0.272 | −0.36 (−1.03–0.31) | 0.296 |
| Intercept | 1.83 (1.53–2.13) | <0.001 | 1.97 (1.58–2.36) | <0.001 | 5.62 (5.26–5.97) | <0.001 |

Table S4. Unstandardized regression coefficients for recommendations to others (participants excluding inattentive or ineligible respondents). Random effects are as follows: between-person standard deviation ($\sigma_u$) = 1.72, residual standard deviation ($\sigma_e$) = 1.42, intraclass correlation coefficient (ICC) = 0.60. Variance inflation factors for covariates, excluding variables used in the regression discontinuity design (i.e., age, pre-policy status, vaccine types, and all associated interaction terms) ≤ 1.56. CI: confidence interval.

| | B (95% CI) | *P* |
|---|---|---|
| Age (centered at 36) [per 10 years] | 0.47 (0.06–0.87) | 0.023 |
| Pre-policy status (ref: post-policy status) | −0.34 (−0.71–0.03) | 0.069 |
| Age [per 10 years] × Pre-policy status | −0.56 (−0.97–−0.15) | 0.008 |
| Physicians (ref: general public) | 1.62 (1.41–1.84) | <0.001 |
| Influenza (ref: measles) | −0.25 (−0.54–0.04) | 0.094 |
| HPV (ref: measles) | −0.45 (−0.74–−0.15) | 0.003 |
| COVID-19 (ref: measles) | −1.55 (−1.84–−1.25) | <0.001 |
| Influenza * Age [per 10 years] | −0.38 (−0.74–−0.03) | 0.035 |
| Influenza * Pre-policy status | 0.31 (−0.02–0.63) | 0.069 |
| Influenza * Age [per 10 years] * Pre-policy status | 0.43 (0.07–0.80) | 0.020 |
| HPV * Age [per 10 years] | −0.54 (−0.90–−0.19) | 0.003 |
| HPV * Pre-policy status | 0.18 (−0.15–0.51) | 0.277 |
| HPV * Age [per 10 years] * Pre-policy status | 0.61 (0.24–0.97) | 0.001 |
| COVID-19 * Age [per 10 years] | −1.32 (−1.68–−0.96) | <0.001 |
| COVID-19 * Pre-policy status | 0.67 (0.34–0.99) | <0.001 |
| COVID-19 * Age [per 10 years] * Pre-policy status | 1.54 (1.17–1.90) | <0.001 |
| Perceived benefits | 0.76 (0.74–0.77) | <0.001 |
| Perceived risks | −0.12 (−0.14–−0.10) | <0.001 |
| Man (ref: woman) | −0.00 (−0.13–0.12) | 0.957 |
| With Children (ref: without children) | 0.17 (0.01–0.32) | 0.032 |
| No answer (ref: without children) | −0.61 (−1.34–0.12) | 0.103 |
| With partner (ref: without partner) | 0.14 (−0.01–0.29) | 0.074 |
| No answer (ref: without partner) | 0.13 (−0.62–0.88) | 0.731 |
| Intercept | 1.60 (1.23–1.97) | <0.001 |

Table S5. Unstandardized regression coefficients for recommendations to others (HPV). Variance inflation factors for covariates, excluding variables used in the regression discontinuity design (i.e., age, pre-policy status, and interaction terms) ≤ 1.52. CI: confidence interval.

| | B (95% CI) | *P* |
|---|---|---|
| Age (centered at 32) [per 10 years] | 0.27 (−0.25–0.79) | 0.316 |
| Pre-policy status (ref: post-policy status) | −0.39 (−0.73–−0.06) | 0.022 |
| Age [per 10 years] × Pre-policy status | −0.32 (−0.85–0.20) | 0.231 |
| Physicians (ref: general public) | 1.51 (1.28–1.73) | <0.001 |
| Perceived benefits | 0.71 (0.69–0.74) | <0.001 |
| Perceived risks | −0.02 (−0.04–0.01) | 0.276 |
| Man (ref: woman) | 0.21 (0.09–0.33) | <0.001 |
| With Children (ref: without children) | 0.19 (0.04–0.33) | 0.013 |
| No answer (ref: without children) | −0.10 (−0.79–0.59) | 0.779 |
| With partner (ref: without partner) | 0.18 (0.03–0.32) | 0.016 |
| No answer (ref: without partner) | 0.26 (−0.33–0.86) | 0.391 |
| Intercept | 1.31 (0.96–1.67) | <0.001 |
